# Blood Biomarkers for Diagnosis & Differential Diagnosis of Alzheimer’s Disease in Real-World Clinical Populations: A Systematic Review

**DOI:** 10.1101/2025.07.06.25330840

**Authors:** Shivani Suresh, Luciana Maffei, Sarah Bauermeister, Vanessa Raymont

## Abstract

**Background:** Gold standard diagnosis of Alzheimer’s disease (AD) relies on invasive, expensive, and non-scalable methods (cerebrospinal fluid lumbar puncture and Amyloid-positron emission tomography). Blood biomarkers (BBMs) present a scalable, accessible, and resource-efficient diagnostic alternative.

**Objective:** To investigate the diagnostic and differential diagnostic performance of three clinically relevant plasma biomarkers: phosphorylated tau-217 (pTau217), glial fibrillary acidic protein (GFAP), and neurofilament light chain (NFL) for biologically confirmed AD patients in real-world, clinical settings.

**Methods:** A systematic search was conducted across 5 databases for peer-reviewed studies between January 2019-January 2025. A narrative synthesis was conducted for eligible studies.

**Results:** 13 studies (n=4686 participants) were included. All studies were cross-sectional, and investigated populations recruited from memory clinics, neurology departments, or clinical cohorts. Diagnostic performance of pTau217 was consistently high (AUC > 0.90 across all comparisons). GFAP and NFL showed moderate and variable accuracy (AUCs ranging from <0.75 to >0.90). No studies assessed combinations of all 3 biomarkers. Methodological and assay heterogeneity was common.

**Conclusion:** Plasma pTau217 demonstrated strong diagnostic accuracy and promise for diagnosis of AD. GFAP and NFL displayed inconsistent results, but could provide complementary information, particularly for differential diagnosis. Further standardized studies in underrepresented populations are required to validate and enable BBM implementation in clinical settings.

## Introduction

Gold standard diagnostic techniques for Alzheimer’s disease (AD), like positron emission tomography (PET) neuroimaging and cerebrospinal fluid (CSF) lumbar puncture, are invasive, expensive and time intensive. Consequently, their use is limited to high-income countries and specialist tertiary-care clinics, but rare in low- and middle-income countries (LMICs) and community-based clinics, which often lack advanced healthcare infrastructure (1). This highlights the need for more cost-effective and scalable alternatives to facilitate timely and accurate diagnoses across diverse clinical settings (2).

Recent advancements in highly sensitive detection methods have enabled accurate detection of relevant biomarkers in peripheral biofluids such as blood. This is a timely development, as the approval of anti-amyloid therapies for AD, aducanumab in 2021 and lecanemab in 2023, has underscored the need for early and accurate diagnoses, and subsequent streamlining of patients into the appropriate clinical trial (3).

The clinical promise of Alzheimer’s disease blood biomarkers is highlighted in the National Institute on Aging-Alzheimer’s Association’s (NIA-AA) most recent guidelines, ‘*Revised criteria for diagnosis and staging of Alzheimer’s disease*’ (4, 5). Updating the 2018 ATN Research Framework (key differences summarized in **Table 1**) (5), this document points to the imminent clinical applicability of blood biomarkers, particularly those that are likely to receive regulatory approval. These are phosphorylated tau-217 (pTau217), glial fibrillary acidic protein (GFAP), and neurofilament light chain (NFL) (4).

**Table 1.** Differences in biomarker categorization between NIA-AA Research Framework (2018) and Clinical Criteria for Staging and Diagnosis (2024). Note: the following biomarkers are included for conceptual purposes but have not undergone the same degree of testing as other Core biomarkers: P-tau231, p-tau205, MTBR-tau243, and non-phosphorylated tau.

|  | Category name | Sub-category | Associated Biomarkers |  |
| --- | --- | --- | --- | --- |
|  |  |  | CSF or Plasma | Imaging |
| 2018 Biomarker Categorization | A | / | CSF Aβ42, or Aβ42/Aβ40 ratio | Amyloid PET |
|  | T | / | CSF phosphorylated tau | Tau PET |
|  | N | / | CSF total tau | Anatomic MRI<br>FDG PET |
| 2024 Biomarker Categorization | A | Core biomarkers: Core 1 | Aβ42 | Amyloid PET |
|  | T1 |  | p-tau 217, p-tau 181, p-tau 231 | / |
|  | T2 | Core biomarkers: Core 2 | pT205, MTBR-243, non-phosphorylated tau fragments | Tau PET |
|  | N | Biomarkers of non-specific processes involved in AD pathophysiology | NFL | Anatomic MR or CT, FDG PET |
|  | I |  | GFAP | / |
|  | V | Biomarkers of non-AD co-pathology | / | Anatomic infarction, WMH |
|  | S |  | CSF αSyn-SAA | / |

**Table 2.** Study Characteristics.

| First Author (Year) | Title | Country | Study Setting | Sample Size | Age (as reported) | %Females (as reported) |
| --- | --- | --- | --- | --- | --- | --- |
| Baiardi (2022) | Diagnostic value of plasma p-tau181, NfL, and GFAP in a clinical setting cohort of prevalent neurodegenerative dementias | Italy | <u>Clinic.</u> Neuropathology Laboratory (NP-Lab) at the Institute of Neurological Science of Bologna, Italy. | 376 | Mean Age (SD): AD = 67.8 (9.3), HC = 61.7 (4.9), FTD = 62.9 (8.9); PSP = 69.2 (10.2); CBS = 71.3 (7.2); DLB = 73.7 | AD = 55.7, HC = 43.3, FTD = 57.6, PSP = 35.5, CBS = 62.1, DLB = 28.6, SNAP = 49.0 |
|  |  |  |  |  | (6.7); SNAP = 66.2 (9.5) |  |
| Cecchetti (2024) | Diagnostic accuracy of automated Lumipulse plasma pTau-217 in Alzheimer's disease: a real-world study | Italy | <u>Clinic.</u> Neurology Unit of IRCCS San Raffaele Scientific Institute | 98 | Mean ± SD (min-max): AD-DEM = 70.9 ± 7.32 (53.30–83.78); AD-MCI = 73.07 ± 6.84 (53.26–84.04); NonAD-DEM = 71.17 ± 7.33 (56.34–80.73); NonAD-MCI = 69.51 ± 8.24 (51.54–83.56) | AD-DEM = 44.4, AD-MCI = 43.5, NonAD-DEM = 60.0, NonAD-MCI = 45.5 |
| Chen (2024) | Diagnostic value of isolated plasma biomarkers and its combination in neurodegenerative dementias: A multicenter cohort study | China | <u>4 Cohorts.</u> Zhejiang University School of Medicine (First Affiliated Hospital, Second Affiliated Hospital, Sir Run Run Shaw Hospital, and Affiliated Zhejiang Hospital) | 112 | Mean ±SD: Total = 64.4 ± 10.0, HC = 65.7 ± 9.1, AD = 62.8 ± 10.7, FTD = 64.3 ± 9.00, PSP = 71.8 ± 5.9 | Overall = 51.7, HC = 50.0, AD = 48.4, FTD = 62.5, PSP = 58.3 |
| Kivisäkk (2023) | Plasma biomarkers for diagnosis of Alzheimer's disease and prediction of cognitive decline in individuals with mild cognitive impairment | USA | <u>Cohort.</u> MADRC-LC study, a longitudinal observational study of cognitive aging, AD, and AD-related disorders. | 238 in diagnostic sample [Total N=307] | Mean ± SD: CN = 72.6 ± 10.4, AD = 74.2 ± 10.6, OND= 69.4 ± 10.9 | CN = 56.7, AD = 47.4, OND = 45.3 |
| Lopes das Neves (2023) | Serum Neurofilament Light Chain in the Diagnostic Evaluation of Patients with Cognitive | Portugal | <u>Clinic.</u> Dementia consultation service. | 365 | CTRs = 65.2 (±10.3); MCI = 67.2 (±9.1); AD = 64.0 (±6.4); FTD = 62.9 (±6.8) years | CTRs = 58.8, MCI data is incorrect in Table S1, AD = 60.5, FTD = 45.9 |
|  | Symptoms in the Neurological Consultation of a Tertiary Center |  |  |  |  |  |
| Oeckl (2019) | Glial Fibrillary Acidic Protein in Serum is Increased in Alzheimer's Disease and Correlates with Cognitive Impairment | Germany | <u>Clinic.</u> Recruitment was done at Ulm University Hospital, Department of Neurology, LMU Munich, TU Munich, and University of Erlangen-Nuremberg. | 127 | median age (IQR): controls = 66 (57–74), PD = 73 (64–76), bvFTD = 64 (56–71), AD = 71 (67–78), DLB/PDD = 74 (70–76) | Overall = 39.4, Controls = 26.5, PD = 18.2, bvFTD = 42.9, AD = 67.9, DLB/PDD = 26.3 |
| Oeckl (2022) | Serum GFAP differentiates Alzheimer's disease from frontotemporal dementia and predicts MCI-to-dementia conversion | Portugal, Netherlands and Germany | <u>Clinic.</u> Patients recruited from University Hospital of Coimbra (Portugal), Radboud University Medical Center (Netherlands), the Technical University of Munich (Germany) and Ulm University Hospital (Germany). | 610 | median age (IQR): controls = 63 (57–69), bvFTD = 62 (57–68); MCI-AD = 71 (64–74), AD dementia = 69 (62–76) | Overall = 54.1, Controls = 48.8, bvFTD = 45.7, MCI-AD = 57.7, AD dementia = 60.4 |
| Palmqvist (2020) | Discriminative accuracy of plasma phospho-tau217 for Alzheimer disease vs other neurodegenerative disorders | Sweden | <u>Cohort.</u> Subjects were recruited from the BioFINDER study. | 699 | median (IQR) age: CU = 66.6 (55.3–76.1), MCI = 72.2 (65.3–75.9), AD dementia = 74.2 (70.4–78.1), other NDDs = 72.4 (64.0–76.5) | CU = 54.8, MCI = 44.9, AD dementia = 52.1, other NDDs = 48.5 |
| Sarto (2023) | Diagnostic Performance and Clinical Applicability of Blood-Based Biomarkers in a | Spain | <u>Clinic.</u> Alzheimer's Disease and Other Cognitive Disorders Unit, | 268 in AD biomarker subcohort [Total N=385] | mean (SD) age: CU = 61.7 (8.2), overall clinical | CU = 78, Overall clinical population = 53, SCD = |
|  | Prospective Memory Clinic Cohort |  | Hospital Clinic, Barcelona, Spain. |  | population = 66.7 (7.5), SCD = 66.9 (5.4), MCI = 67.1 (7.4), AD dementia = 66.2 (6), LBD = 68.5 (6.7), FTD = 66.2 (9.4) | 62, MCI = 55, AD dementia = 62, LBD = 30, FTD = 46 |
| Shen (2023) | Plasma Glial Fibrillary Acidic Protein in the Alzheimer Disease Continuum: Relationship to Other Biomarkers, Differential Diagnosis, and Prediction of Clinical Progression | China | <u>Clinic and cohort.</u> Subjects recruited from the Memory Clinic of the Huashan Hospital of Fudan University (Shanghai) and the Chinese Alzheimer Biomarker and Lifestyle (CABLE) study | 700 | mean (SD) age: CN = 60 (9), MCI = 64 (11), AD = 59 (8), PCA = 56 (5), PPA = 61 (8), bvFTD = 59 (9), FTD-MND = 61 (7), PSP = 66 (7), VaD = 65 (8), DLB = 68 (9), MSA = 59 (8), PD = 64 (10), SCA = 46 (12), ALS = 51 (20) | CN = 55.9, MCI = 51.8, AD = 59.6, PCA = 53.8, PPA = 62.8, bvFTD = 50.0, FTD-MND = 71.4, PSP = 41.2, VaD = 28.9, DLB = 46.4, MSA = 44, PD = 45, SCA = 50.0, ALS = 50.0 |
| Thijssen (2021) | Association of Plasma P-tau217 and P tau181 with clinical phenotype, neuropathology, and imaging markers in Alzheimer's disease and frontotemporal lobar degeneration: a retrospective diagnostic performance study | USA | <u>Clinic.</u> Recruitment was done from the University of California San Francisco (UCSF) Memory and Aging Center, an NIA ADRC, and from the Advancing Research and Treatment for Frontotemporal Lobar Degeneration (ARTFL) consortium | 593 | Mean age (SD): Overall = 64.3 (13), CN = 60.9 (18), MCI = 65.5 (13), ADclin = 65.3 (10), lvPPA = 63.1 (9), PCA = 57.6 (11), CBS = 67.3 (8), PSP = 68.5 (7), bvFTD = 61.2 (10), nvPPA = 69.8 (7), svPPA = 70.0 (7), DLB = 69.3 (6), TES = 63.2 (13) | Overall = 49.7, CN = 53.4, MCI = 44.4, ADclin = 56.9, lvPPA = 53.3, PCA = 100 (n=2), CBS = 54.4, PSP = 54.1, bvFTD = 40.3, nvPPA = 46.9, svPPA = 59.3, DLB = 35.7, TES = 0 (n=13) |
| Thijssen (2022) | Differential diagnostic performance of a panel of plasma biomarkers for different types of dementia | Netherlands | <u>Cohort.</u> Patients from both Cohort 1 and 2 were recruited from the Amsterdam Dementia Cohort. | 160 (Cohort 1) +152 (Cohort 2) | Median age (IQR): Cohort 1: Controls = 56 (53–59), AD = 58 (55–59), FTD = 64 (61–70), DLB = 70 (62–74), Overall = 59 (56–66); Cohort 2: Controls = 63 (59–66), AD = 63 (59–67), FTD = 63 (59–67), DLB = 67 (64–69), Overall = 64 (60–68) | Cohort 1: Controls = 50, AD = 50, FTD = 55, DLB = 30, Overall = 46; Cohort 2: Controls = 53, AD = 53, FTD = 53, DLB = 8, Overall = 41 |
| Vrillon (2023) | Comparison of CSF and plasma NfL and pNfH for Alzheimer's disease diagnosis: a memory clinic study | France | <u>Clinic.</u> Patients recruited from a tertiary memory center - the Center of Cognitive Neurology, at the University Hospital Lariboisière, Paris. | 188 | Mean age (SD): Controls = 63.4 (10.1), non-AD MCI = 68.7 (9.7), AD-MCI = 72.1 (7.4), AD dementia = 71.8 (8.4), non AD dementia = 67.4 (7.6) | Controls = 68, Non-AD MCI = 66, AD-MCI = 53, AD dementia = 64, non-AD dementia = 54 |

**Table 3.**
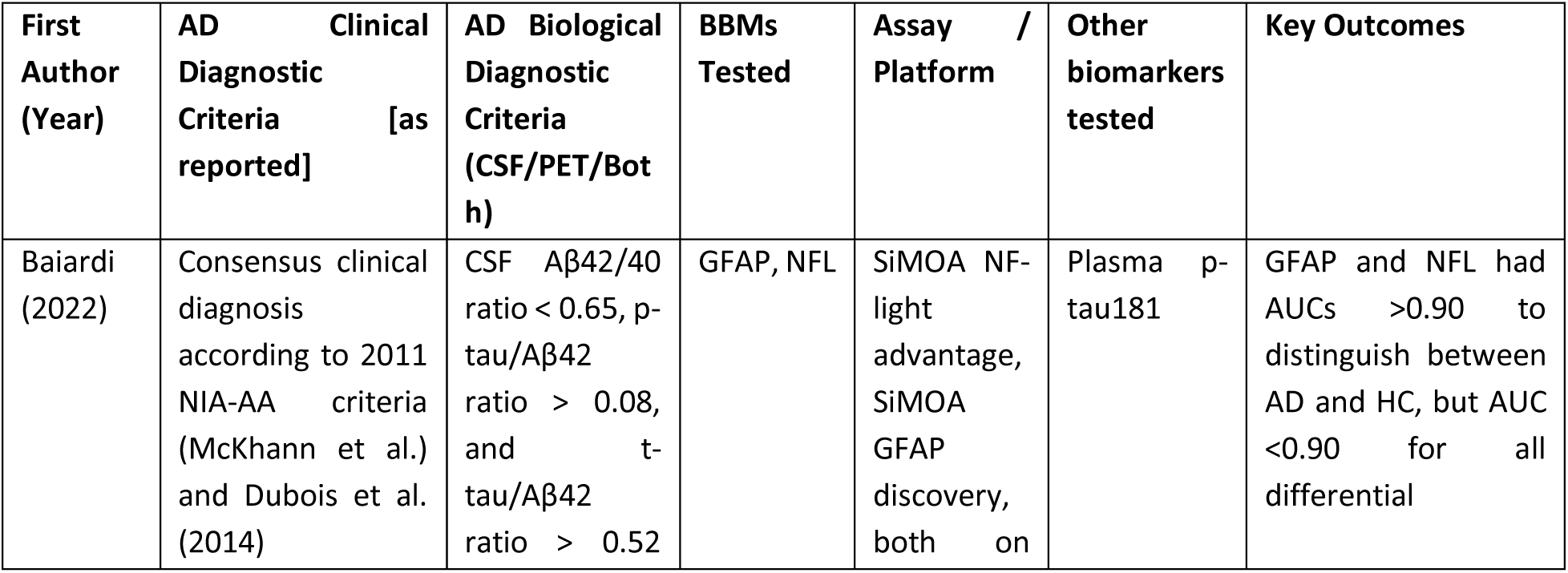
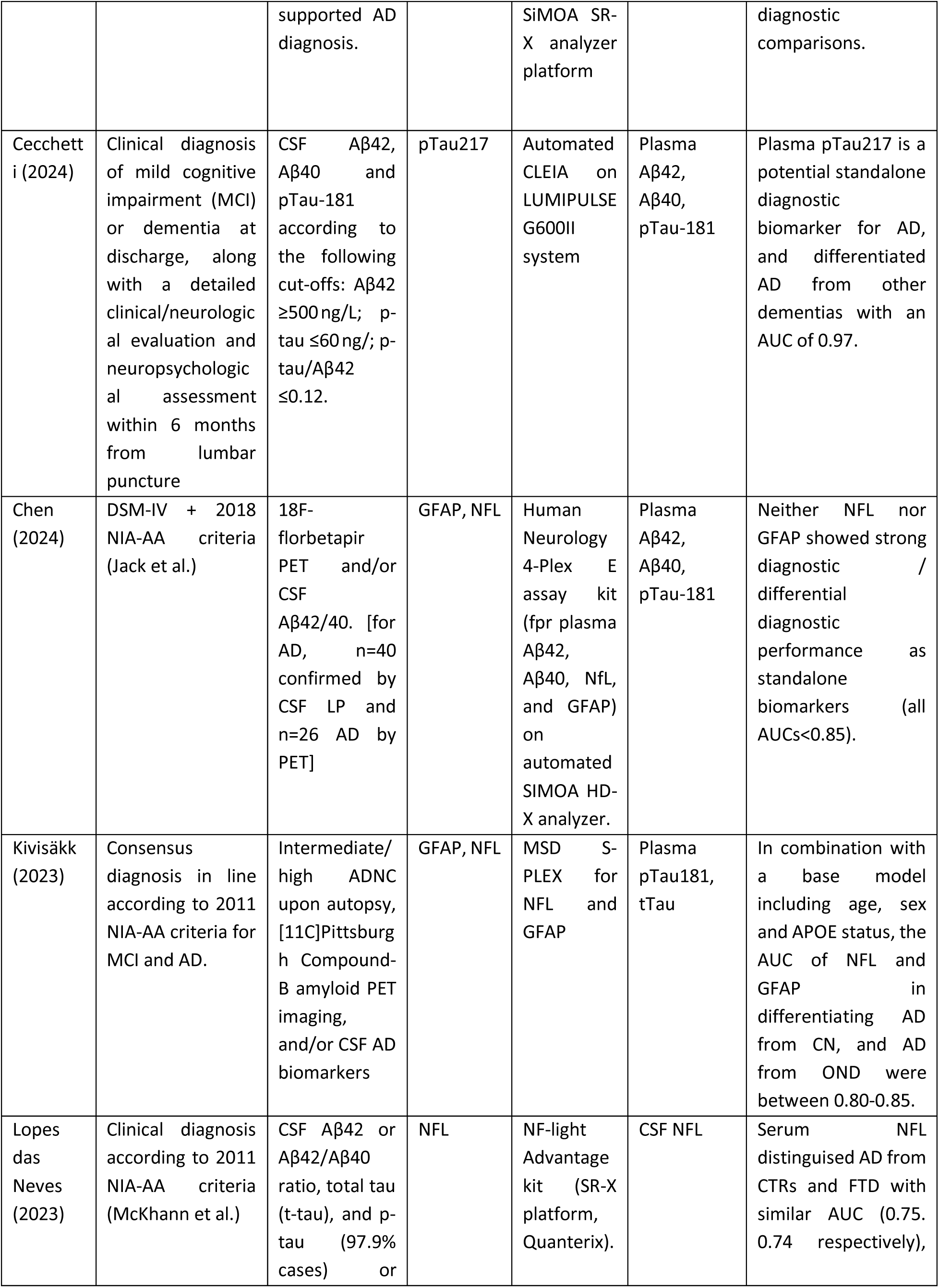
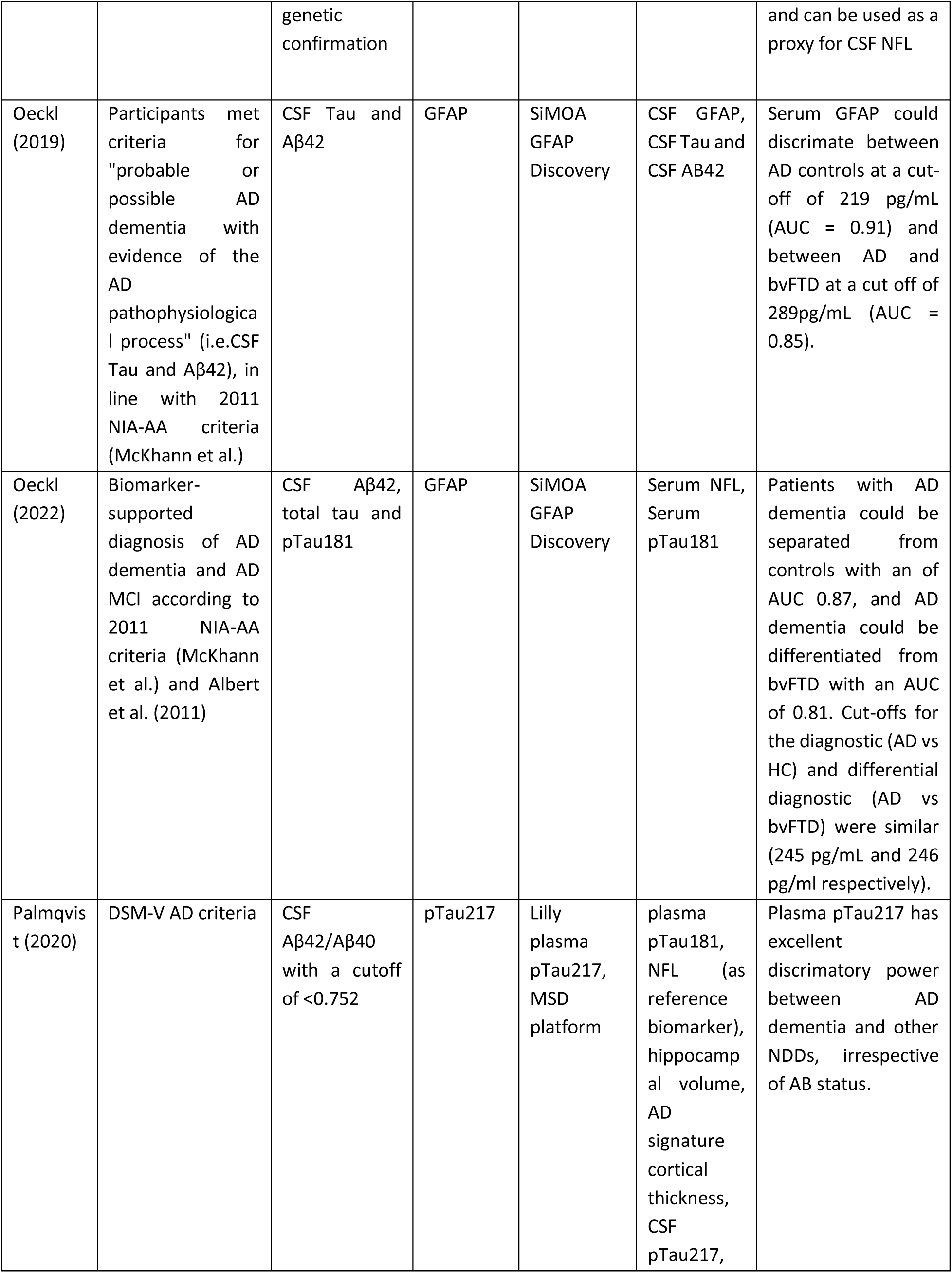
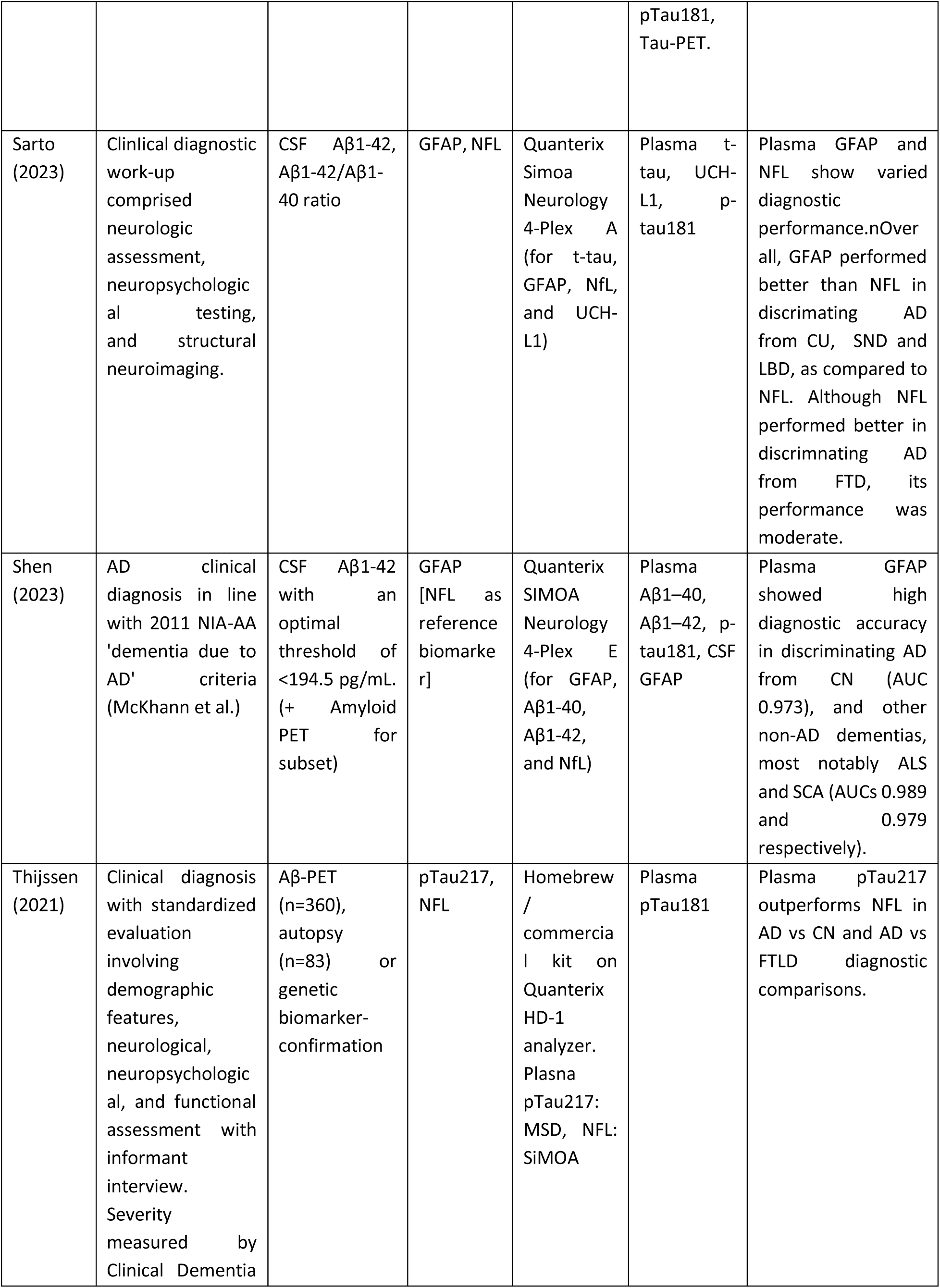
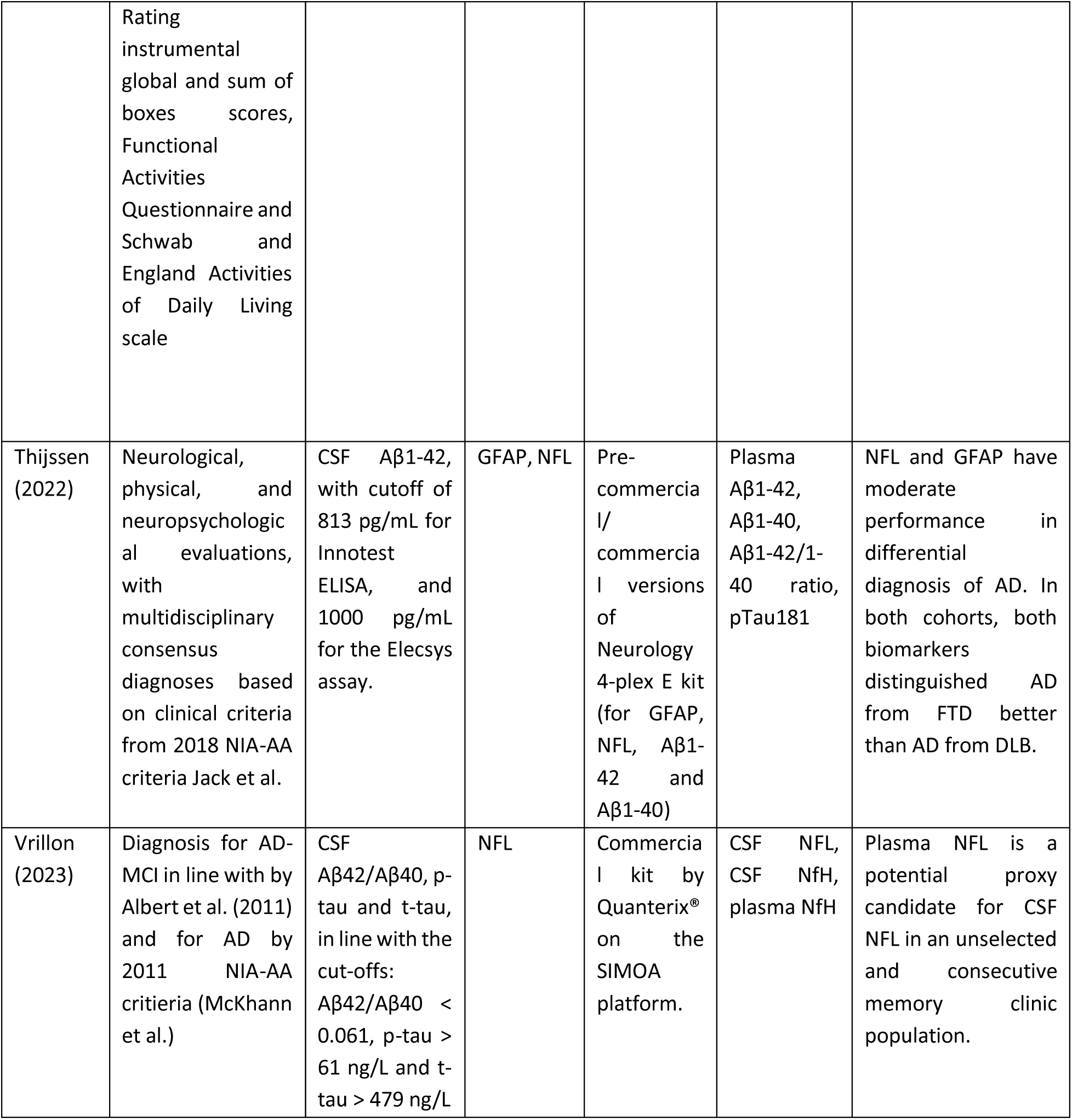
Diagnostic Criteria and Blood Biomarkers.

| First Author (Year) | AD Clinical Diagnostic Criteria [as reported] | AD Biological Diagnostic Criteria (CSF/PET/Both) | BBMs Tested | Assay / Platform | Other biomarkers tested | Key Outcomes |
| --- | --- | --- | --- | --- | --- | --- |
| Baiardi (2022) | Consensus clinical diagnosis according to 2011 NIA-AA criteria (McKhann et al.) and Dubois et al. (2014) | CSF Aβ42/40 ratio < 0.65, p-tau/Aβ42 ratio > 0.08, and t-tau/Aβ42 ratio > 0.52 | GFAP, NFL | SiMOA NF-light advantage, SiMOA GFAP discovery, both on | Plasma p-tau181 | GFAP and NFL had AUCs >0.90 to distinguish between AD and HC, but AUC <0.90 for all differential |
|  |  | supported AD diagnosis. |  | SiMOA SR-X analyzer platform |  | diagnostic comparisons. |
| Cecchetti (2024) | Clinical diagnosis of mild cognitive impairment (MCI) or dementia at discharge, along with a detailed clinical/neurological evaluation and neuropsychological assessment within 6 months from lumbar puncture | CSF Aβ42, Aβ40 and pTau-181 according to the following cut-offs: Aβ42 ≥500ng/L; p-tau ≤60ng/L; p-tau/Aβ42 ≤0.12. | pTau217 | Automated CLEIA on LUMIPULSE G600II system | Plasma Aβ42, Aβ40, pTau-181 | Plasma pTau217 is a potential standalone diagnostic biomarker for AD, and differentiated AD from other dementias with an AUC of 0.97. |
| Chen (2024) | DSM-IV + 2018 NIA-AA criteria (Jack et al.) | 18F-florbetapir PET and/or CSF Aβ42/40. [for AD, n=40 confirmed by CSF LP and n=26 AD by PET] | GFAP, NFL | Human Neurology 4-Plex E assay kit (for plasma Aβ42, Aβ40, NFL, and GFAP) on automated SIMOA HD-X analyzer. | Plasma Aβ42, Aβ40, pTau-181 | Neither NFL nor GFAP showed strong diagnostic / differential diagnostic performance as standalone biomarkers (all AUCs<0.85). |
| Kivisäkk (2023) | Consensus diagnosis in line according to 2011 NIA-AA criteria for MCI and AD. | Intermediate/high ADNC upon autopsy, [11C]Pittsburgh Compound-B amyloid PET imaging, and/or CSF AD biomarkers | GFAP, NFL | MSD S-PLEX for NFL and GFAP | Plasma pTau181, tTau | In combination with a base model including age, sex and APOE status, the AUC of NFL and GFAP in differentiating AD from CN, and AD from OND were between 0.80-0.85. |
| Lopes das Neves (2023) | Clinical diagnosis according to 2011 NIA-AA criteria (McKhann et al.) | CSF Aβ42 or Aβ42/Aβ40 ratio, total tau (t-tau), and p-tau (97.9% cases) or | NFL | NF-light Advantage kit (SR-X platform, Quanterix). | CSF NFL | Serum NFL distinguished AD from CTRs and FTD with similar AUC (0.75. 0.74 respectively), |
|  |  | genetic confirmation |  |  |  | and can be used as a proxy for CSF NFL |
| Oeckl (2019) | Participants met criteria for "probable or possible AD dementia with evidence of the AD pathophysiological process" (i.e.CSF Tau and Aβ42), in line with 2011 NIA-AA criteria (McKhann et al.) | CSF Tau and Aβ42 | GFAP | SiMOA GFAP Discovery | CSF GFAP, CSF Tau and CSF AB42 | Serum GFAP could discriminate between AD controls at a cut-off of 219 pg/mL (AUC = 0.91) and between AD and bvFTD at a cut off of 289pg/mL (AUC = 0.85). |
| Oeckl (2022) | Biomarker-supported diagnosis of AD dementia and AD MCI according to 2011 NIA-AA criteria (McKhann et al.) and Albert et al. (2011) | CSF Aβ42, total tau and pTau181 | GFAP | SiMOA GFAP Discovery | Serum NFL, Serum pTau181 | Patients with AD dementia could be separated from controls with an of AUC 0.87, and AD dementia could be differentiated from bvFTD with an AUC of 0.81. Cut-offs for the diagnostic (AD vs HC) and differential diagnostic (AD vs bvFTD) were similar (245 pg/mL and 246 pg/ml respectively). |
| Palmqvist (2020) | DSM-V AD criteria | CSF Aβ42/Aβ40 with a cutoff of <0.752 | pTau217 | Lilly plasma pTau217, MSD platform | plasma pTau181, NFL (as reference biomarker), hippocampal volume, AD signature cortical thickness, CSF pTau217, pTau181,<br>Tau-PET. | Plasma pTau217 has excellent discriminatory power between AD dementia and other NDDs, irrespective of AB status. |
| Sarto<br>(2023) | Clinical diagnostic work-up comprised neurologic assessment, neuropsychological testing, and structural neuroimaging. | CSF Aβ1-42, Aβ1-42/Aβ1-40 ratio | GFAP, NFL | Quanterix Simoa Neurology 4-Plex A (for t-tau, GFAP, NFL, and UCH-L1) | Plasma t-tau, UCH-L1, p-tau181 | Plasma GFAP and NFL show varied diagnostic performance. Overall, GFAP performed better than NFL in discriminating AD from CU, SND and LBD, as compared to NFL. Although NFL performed better in discriminating AD from FTD, its performance was moderate. |
| Shen<br>(2023) | AD clinical diagnosis in line with 2011 NIA-AA 'dementia due to AD' criteria (McKhann et al.) | CSF Aβ1-42 with an optimal threshold of <194.5 pg/mL. (+ Amyloid PET for subset) | GFAP [NFL as reference biomarker] | Quanterix SIMOA Neurology 4-Plex E (for GFAP, Aβ1-40, Aβ1-42, and NFL) | Plasma Aβ1-40, Aβ1-42, p-tau181, CSF GFAP | Plasma GFAP showed high diagnostic accuracy in discriminating AD from CN (AUC 0.973), and other non-AD dementias, most notably ALS and SCA (AUCs 0.989 and 0.979 respectively). |
| Thijssen<br>(2021) | Clinical diagnosis with standardized evaluation involving demographic features, neurological, neuropsychological, and functional assessment with informant interview. Severity measured by Clinical Dementia | Aβ-PET (n=360), autopsy (n=83) or genetic biomarker-confirmation | pTau217, NFL | Homebrew / commercial kit on Quanterix HD-1 analyzer. Plasma pTau217: MSD, NFL: SIMOA | Plasma pTau181 | Plasma pTau217 outperforms NFL in AD vs CN and AD vs FTLD diagnostic comparisons. |
|  | Rating instrumental global and sum of boxes scores, Functional Activities Questionnaire and Schwab and England Activities of Daily Living scale |  |  |  |  |  |
| Thijssen (2022) | Neurological, physical, and neuropsychological evaluations, with multidisciplinary consensus diagnoses based on clinical criteria from 2018 NIA-AA criteria Jack et al. | CSF Aβ1-42, with cutoff of 813 pg/mL for Innotech ELISA, and 1000 pg/mL for the Elecsys assay. | GFAP, NFL | Pre-commercial/ commercial versions of Neurology 4-plex E kit (for GFAP, NFL, Aβ1-42 and Aβ1-40) | Plasma Aβ1-42, Aβ1-40, Aβ1-42/1-40 ratio, pTau181 | NFL and GFAP have moderate performance in differential diagnosis of AD. In both cohorts, both biomarkers distinguished AD from FTD better than AD from DLB. |
| Vrillon (2023) | Diagnosis for AD-MCI in line with by Albert et al. (2011) and for AD by 2011 NIA-AA criteria (McKhann et al.) | CSF Aβ42/Aβ40, p-tau and t-tau, in line with the cut-offs: Aβ42/Aβ40 < 0.061, p-tau > 61 ng/L and t-tau > 479 ng/L | NFL | Commercial kit by Quanterix® on the SIMOA platform. | CSF NFL, CSF NfH, plasma NfH | Plasma NFL is a potential proxy candidate for CSF NFL in an unselected and consecutive memory clinic population. |

**Table 4.**
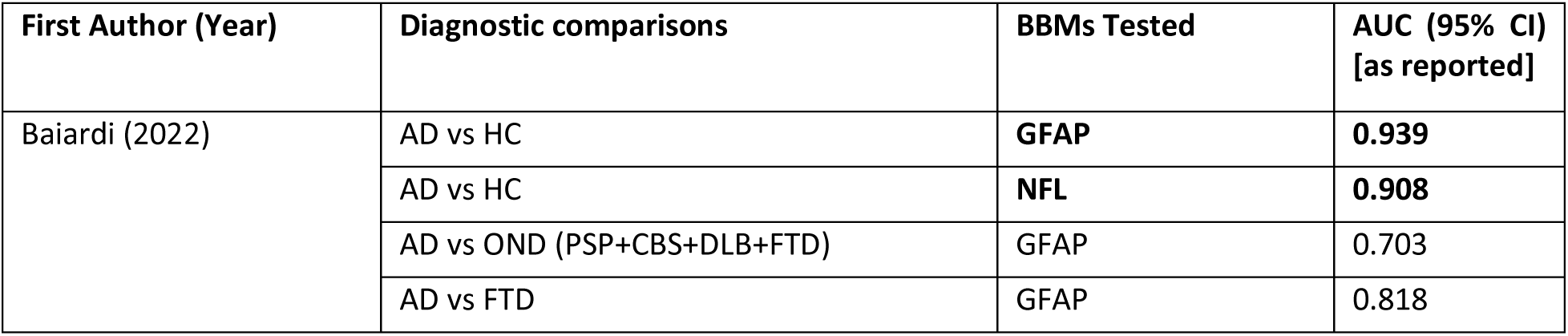
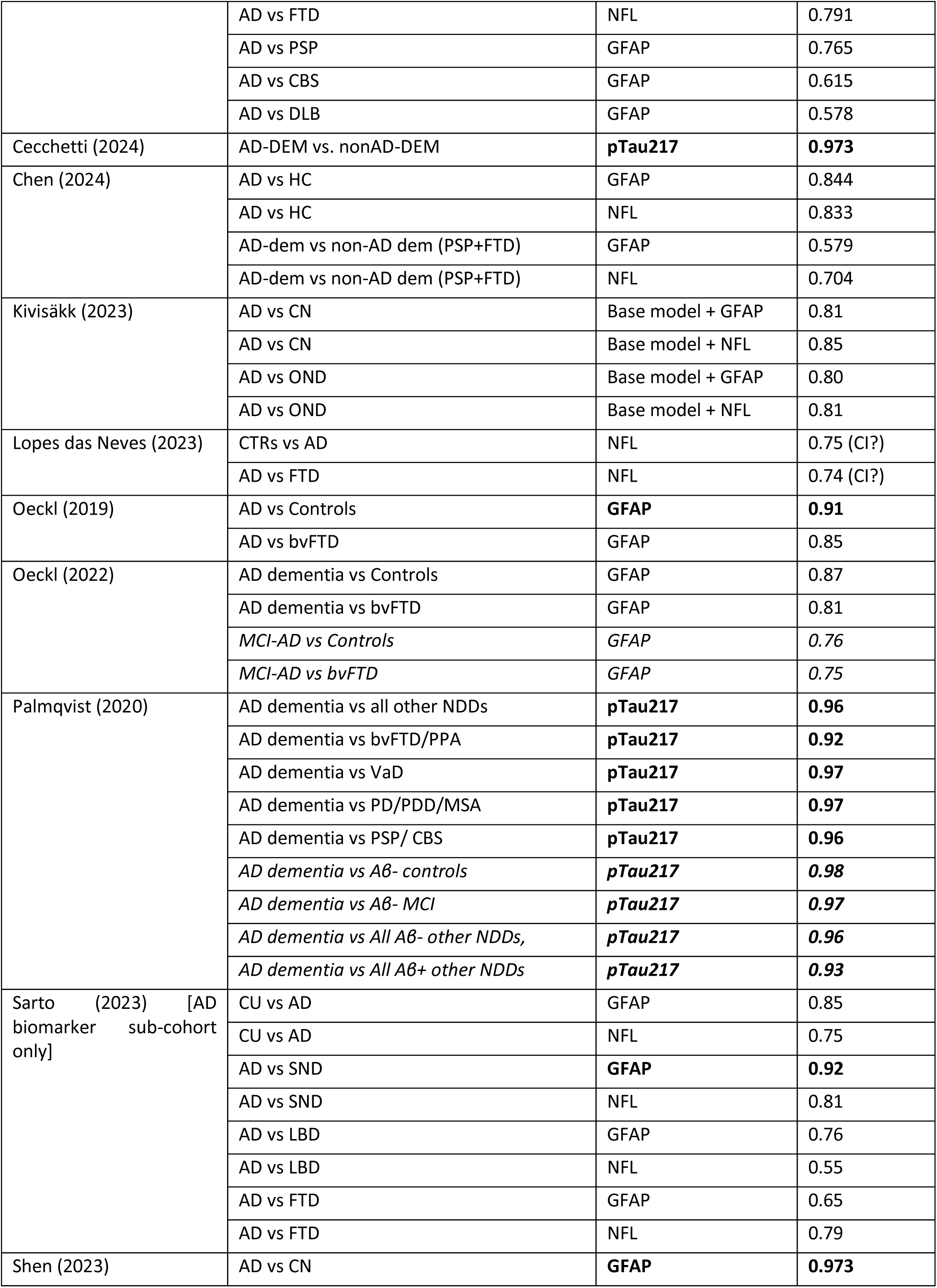
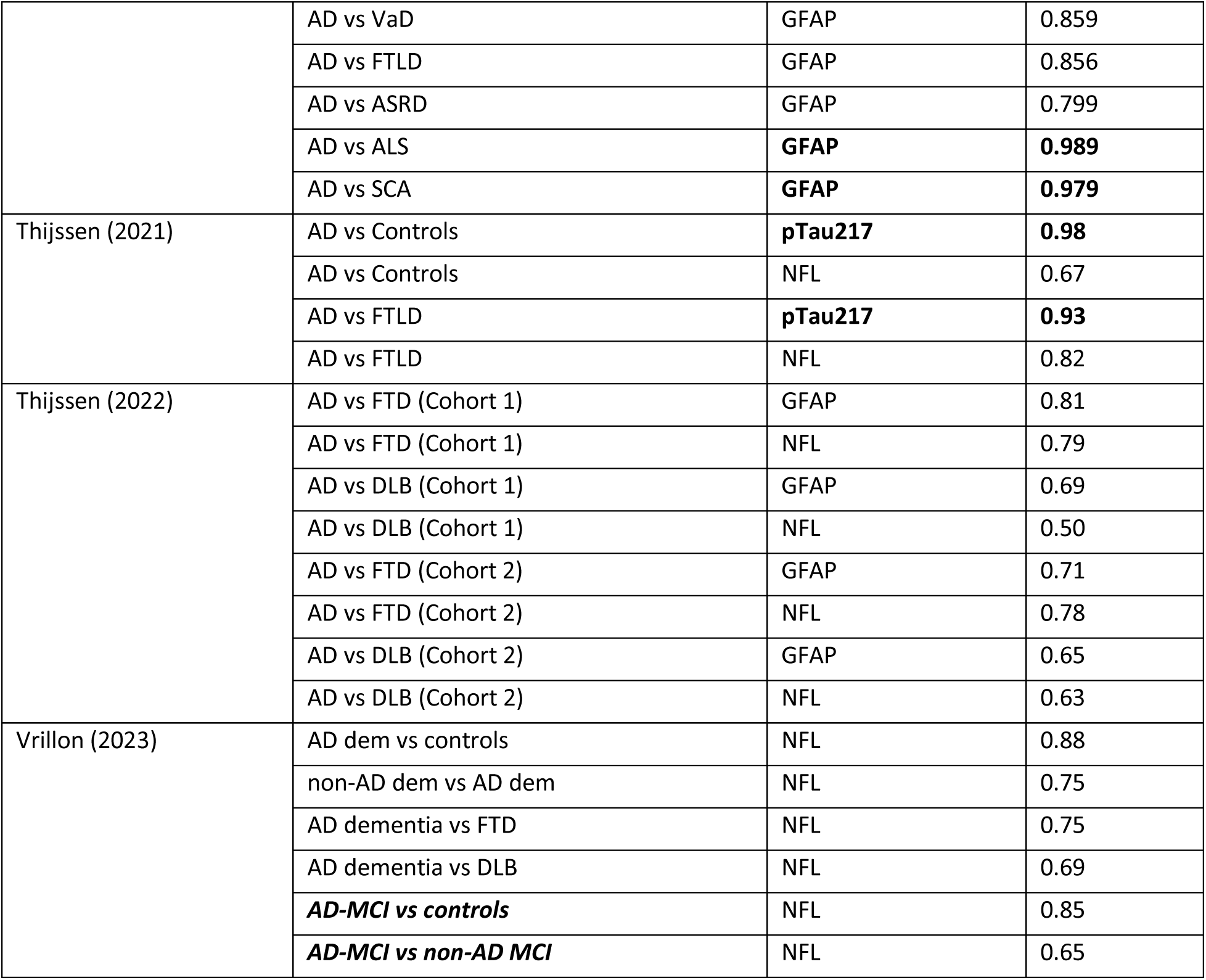
Outcomes.

Current evidence suggests that these biomarkers are clinically relevant: plasma pTau217 has shown diagnostic performance superior to other phosphorylated tau isoforms in both preclinical and clinical stages of AD dementia (6–8). Additionally, both GFAP and NFL are known to be elevated in AD brains (9), and CSF (10–13), and have been shown to differentiate AD from other dementias (14–16). Further, blood biomarkers have arguably the most utility as screening and diagnostic tools in clinical settings, where patients present with a variety of co-pathologies. In such contexts, a combination of AD-specific biomarkers like pTau217, and non-specific biomarkers of associated neurodegenerative and inflammatory pathological processes (NFL and GFAP, respectively), could prove to have immense clinical significance.

Considering the above, this systematic review evaluates the diagnostic and differential diagnostic utility of pTau217, GFAP and NFL in Alzheimer’s disease. Specifically, this review focuses on studies conducted in real-world clinical populations (e.g. memory clinics), where blood biomarker roll-out is most likely, and will offer an accessible alternative to present diagnostic techniques, that can be scaled to diverse, under-resourced settings including LMICs.

## Methods

### Search & Screening

The systematic review was undertaken in accordance with the PRISMA guidelines **[source]**, designed according to the PICOS framework (**figure 1**), and was registered on the International Prospective Register of Systematic Reviews (PROSPERO: https://www.crd.york.ac.uk/PROSPERO/view/CRD42024524962/), registration number CRD42024524962.

**Figure 1.**
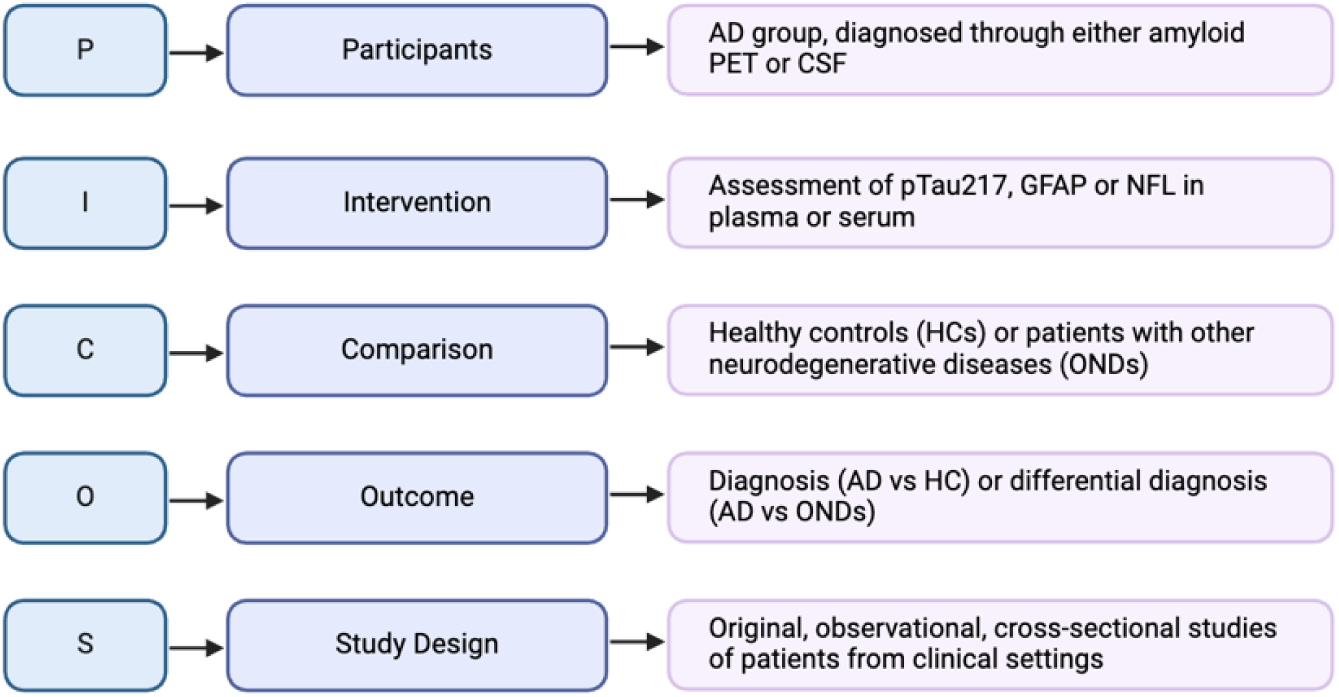
Review design according to PICOS framework.

The systematic search was carried out in two phases, an initial search for publications between 1^st^ January 2019, and a second, update search from 31^st^ December 2023 to 27^th^ January 2025. Both searches included publications across multiple databases (PubMed, EMBASE, Global Health, Journals@OVID and PsycINFO), and were limited to articles published in English in peer-reviewed journals.

Articles were selected as per the specific inclusion criteria: (1) the outcome of the article was to assess diagnostic or differential diagnostic ability of a biomarker of interest, or any combinations of the biomarkers of interest, (2) the population was partially or completely recruited from a clinical setting (for instance, a memory clinic or neurology department in a hospital), (3) the study had a clearly defined Alzheimer’s disease patient group, in which participants had both a clinical diagnosis consistent with AD (i.e., etiological confirmation) and biological confirmation of AD pathology via CSF or PET biomarkers.

Articles were excluded if (1) the study design and outcomes were in line with risk prediction or prognosis only, without any diagnostic element, (2) the population was exclusively cognitively normal, consisted of individuals with memory complaints without an AD-confirmed group, or was subject to strict recruitment criteria (for instance, a cohort study which categorically excluded patients with AD or other dementia diagnoses), (3) the population represented genetic variants of AD alone, (4) the biomarker of interest was measured in combination with a biomarker other than GFAP, NFL or pTau217, or was measured using a non-scalable method (5) the biomarker of interest was used as a reference, to assess efficacy of another biomarker or an intervention, (6) the article was associative, or aimed at elucidating underlying mechanisms of disease, neuropathology, (7) the study was carried out in a biofluid other than blood, or not in a human population (e.g. cell lines, mice), (8) any grey literature (conference abstract, poster presentation, book chapter, theses, etc), meta-analyses or reviews.

Search results were imported into Rayyan, and duplicates were removed. Title-and abstract-screening, and subsequent full text screening were done independently by SS and LM. Any disagreements during the screening process were resolved by a consensus between SS, LM, SB and VR.

### Quality assessment & Data extraction

Quality assessment of all included articles was conducted using the QADAS-2 tool by SS and LM. Signaling questions in all 4 domains of the QADAS questionnaire (patient selection, index test, reference standard and flow and timing) were tailored to ensure review-specific assessment of bias. Relevant data on study characteristics, biomarker- and diagnostic information, and outcomes were extracted by SS and LM. Narrative synthesis was conducted by SS.

## Results

### Study selection

The search identified 4272 articles across five databases (PubMed, Journals @ OVID, Embase, Global Health, and PsycINFO) (**Figure 2**). After eliminating 2296 duplicates, 1976 articles remained for title- and abstract screening. At this stage, 1908 articles were excluded as they were irrelevant to the review question, leaving 68 articles for full-text review. Of these, the full-text version of 2 reports was not available, and 49 reports were excluded for having ineligible diagnostic measures, biomarkers, outcomes, patient populations, or study design, and 4 were excluded as they were poster presentations or conference abstracts. Consequently, 13 articles were included in the review (17–29).

**Figure 2.**
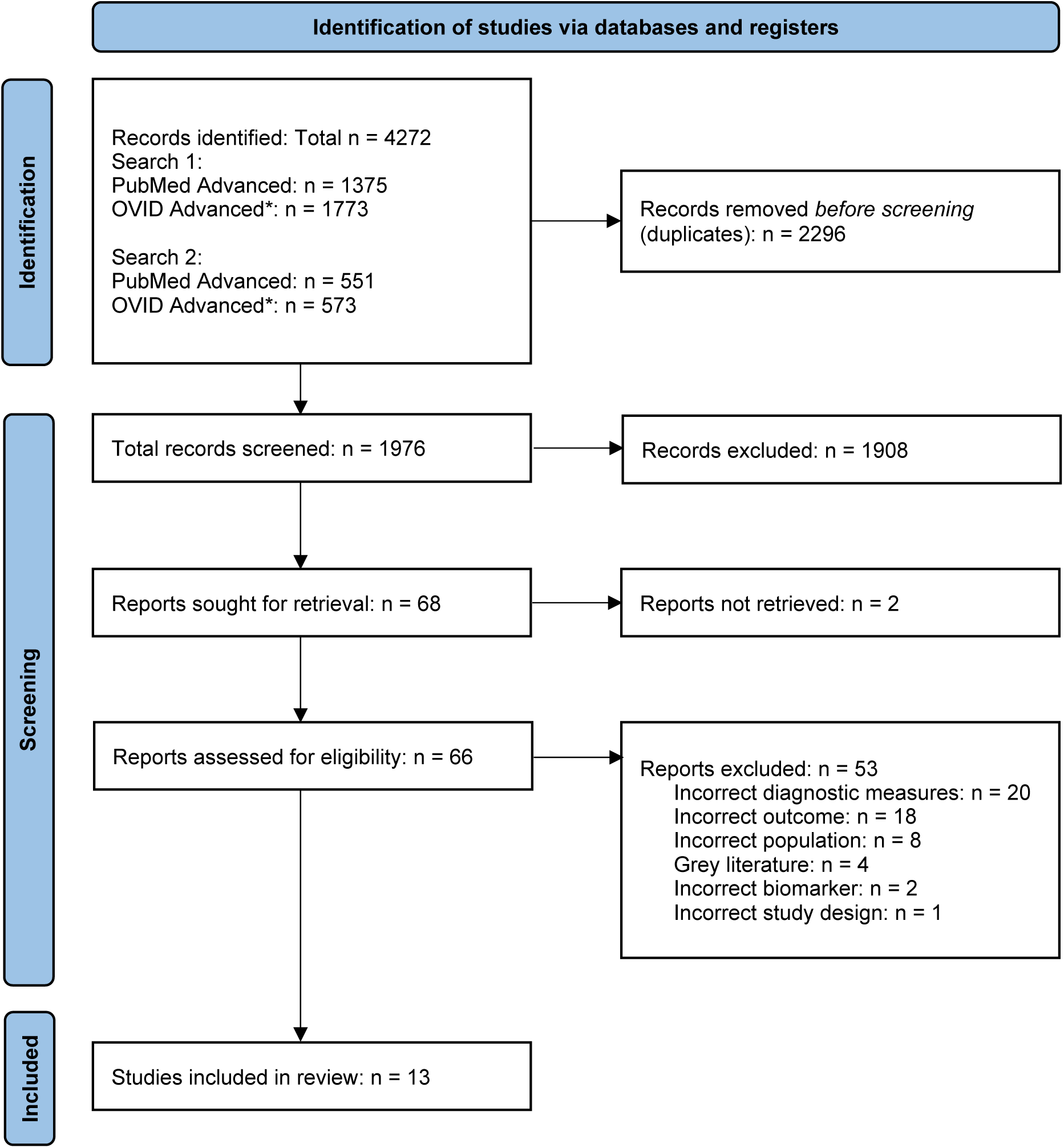
PRISMA flowchart for Systematic Review search and screening. *OVID advanced search included the following databases: Journals @ Ovid, Embase, Global health, PsycINFO

### Study characteristics

Of the thirteen included studies, eight studies were conducted directly in specialist memory clinics or hospital neurology departments, 4 analyzed data from clinical cohorts, and 1 included patient data collected partially from cohorts, and partly from clinic. Four studies explicitly mentioned consecutive patient recruitment, enhancing representation of real-world clinical settings by minimizing selection bias.

A total of 4686 participants were studied across all included articles. In some studies, only specific sub-cohorts relevant to the research question were assessed. Sample sizes were varied across studies, ranging from 98 in Cecchetti et al. (2024) to 700 in Shen et al. (2023). The average/ median age of the AD group was higher than or equal to that of the cognitively unimpaired (CU) group in nine cases and lower in three cases. One study (Ceccheti et al. 2024) lacked a CU group. All studies had a fair representation of both males and females, assessed differences in age between diagnostic groups, and controlled for these factors in analyses if significant effects were found. There was some variability in demographic reporting: 8 studies recorded age as mean (SD) while 4 studies reported median (IQR).

The thirteen studies that met the inclusion criteria of the review represented nine countries. Three studies reported ethnicity: Kivisakk et al. (2023) reported 93.7% non-Hispanic whites, Thijssen et al. (2021) reported an 85% white population, and Shen et al. (2023) reported exclusively Chinese participants.

### Diagnostic Criteria and Blood Biomarkers

There was substantial variability in the methods used to confirm AD diagnosis was observed, both clinically and biologically. Six studies established clinical etiology according to the 2011 NIA-AA criteria, two according to the 2018 NIA-AA criteria, two studies used the DSM criteria, and three studies did not mention any specific guidelines. Despite 11 of 13 selected studies using CSF biomarkers to confirm AD diagnosis, the specific biomarkers varied ranged from only amyloid markers (CSF Aβ1-42, Aβ1-42/Aβ1-40 ratio) to a combination of amyloid and tau markers (such as CSF total tau, p-tau, p-tau181), and hybrid ratios (Aβ42/p-tau ratio). Amyloid PET was utilized in four studies, and genetic confirmation, and pathological diagnosis on autopsy were employed in addition to CSF/PET biomarkers in some cases.

Six studies evaluated more than one blood biomarker of interest, and no study evaluated all three biomarkers. All selected articles assessed other biomarkers, beyond the scope of this review. These were most commonly plasma pTau181 and amyloid markers in both plasma and CSF.

The diagnostic performance of pTau217 was assessed in three studies, all using different assays and platforms for analysis (Automated CLEIA on LUMIPULSE G600II system, Lilly plasma pTau217 on the MSD platform, commercial/ homebrew kit on the Quanterix HD-1 analyser). Along a similar vein, the eight studies that measured GFAP and NFL also used various assays for biomarker measurement. Individual measurements of NFL were either reported using the NF-light advantage kit, or a ‘commerical kit by Quanterix’ (Vrillon et al. 2023), or a ‘homebrew or commercially available kit’ (Thijssen et al. 2021). Additionally, four studies used multiplex assays for the simultaneous measurement of plasma GFAP and NFL. Kivisakk et al. (2023) used an ultrasensitive prototype S-PLEX assay on the MSD platform, Thijssen et al. (2022) and Shen et al. (2023) used the Neurology 4-plex E kit to for simultaneous measurement of GFAP, NFL, Aβ1-42 and Aβ1-40, and Sarto et al. (2023) used the Neurology 4-plex A kit to analyze GFAP, NFL, UCH-L1 and t-tau.

Overall, several assays were used to measure all biomarkers on both MSD and SiMOA platforms. Different versions of the SiMOA platform were used in different studies (e.g. SR-X, HD-1), leading to less standardization whilst measuring the same biomarker. Taken together, this reflects heterogeneity in biomarker measurement techniques.

No study measured the biomarkers relevant to the review in a panel with one another. Combinations of biomarkers of interest with those that are not within the scope of this review were excluded to maintain consistency in the relevant biomarkers.

### Outcomes

Eleven out of the selected thirteen studies reported both diagnostic (AD vs. CU) and differential diagnostic (AD vs. other neurodegenerative disease(s)) comparisons, two reported differential diagnosis alone. Studies whose outcome was solely the detection of amyloid positivity were systematically excluded from this review to maintain focus on the dementia phase of the AD continuum.

Of the three biomarkers of interest, only pTau217 consistently had an accuracy of >0.90 across all studies, and all twelve diagnostic comparisons. Overall, performance of both GFAP and NFL was variable and moderate. Of the thirty diagnostic comparisons in which GFAP was evaluated, six had an AUC > 0.90, sixteen had an AUC >0.75-0.90, and eight reported AUC < 0.75. Twenty four comparisons were investigated for NFL, of which only one had an AUC > 0.90.

## Discussion

This systematic review evaluates the utility of three blood-based biomarkers – phosphorylated tau 217 (p-tau217), glial fibrillary acidic protein (GFAP) and neurofilament light chain protein (NfL), in the diagnosis and differential diagnosis of AD dementia in patients recruited from clinical settings, such as memory clinics and hospital neurology departments. The development of highly sensitive blood-based tests to detect and diagnose Alzheimer’s disease presents a highly resource-efficient, non-invasive and accessible alternative to traditional CSF and PET neuroimaging techniques, that are expensive and non-scalable (30). These biomarkers were recognised in the Alzheimer’s Association Workgroup’s recent ‘Revised Criteria for Diagnosis and Staging of Alzheimer’s disease’, which emphasized their contribution in recent AD dementia research, and highlighted the likelihood of their regulatory approval imminently (4). The revised criteria will play an important role in laying the groundwork for rollout of commercial plasma biomarker tests in coming years, particularly in clinical services, given their likely upscale given the advent of disease modifying therapies (31, 32). In light of this, the current review is timely in synthesising the current literature, pointing out gaps in the literature, and laying the groundwork for future studies.

One of the key recommendations of the Alzheimer’s Association’s 2024 *Revised Criteria* demonstrate an accuracy of >90%, or equivalent to its CSF counterpart. All studies assessing diagnostic utility of plasma p-tau217 are consistent with this recommendation. One of these studies, conducted by Thijssen et al. (2021), also demonstrated the superior performance of p-tau217 as compared to NFL across all diagnostic comparisons. This is expected as NFL is a non-specific biomarker of neurodegeneration, especially in disorders of cognitive dysfunction (12, 33, 34). While the combined performance of p-tau217 and NFL was not explored, individual evaluation of NFL was investigated. Although two independent evaluations of NFL reported moderate diagnostic accuracy, both demonstrated that the diagnostic performance of plasma NfL was equivalent to its CSF counterpart, suggesting increased clinical utility and scalability (26, 27). Similar findings have been reported in longitudinal studies of a cohort of non-demented participants (cognitively normal, or with MCI) (35). However, a meta-analysis evaluating the overall pooled correlation coefficient estimate between NFL in both these biofluids reported only moderate correlations (36).

In contrast to NFL, GFAP showed >90% accuracy in AD diagnosis (Oeckl et al. 2019, Shen et al. 2023) and differential diagnosis, specifically against non-AD pathology (Sarto et al. 2023), as well as amyotrophic lateral sclerosis and spinocerebellar ataxia (Shen et al. 2023). Notably, the study by Shen et al. (2023) is the only selected study to report findings from an LMIC, and could provide valuable insights into the global applicability of GFAP in clinical settings. In the four studies that assessed accuracy of both NFL and GFAP, GFAP outperformed NFL in most diagnostic comparisons. Analogously, GFAP outperformed NFL in predicting AD incidence in population-based community cohort representing AD and other neurodegenerative disorders followed for 17 years, suggesting similar trends in studies aimed at risk prediction and prognosis (37).

The major strengths of this review are the specific focus on clinical populations, evaluation of biomarkers of high clinical relevance, and the validation of blood biomarkers against gold-standard diagnostic methods (PET or CSF). However, some of the initial hypotheses of the review were not met. For instance, combinations or panels of the selected biomarkers only were not performed in any of the included studies. This is surprising for a few reasons. First, it was anticipated that the inclusion of differential diagnosis as an outcome would prompt the selection of articles that looked at AD-specific (p-tau217), along with non-specific (GFAP and NFL) biomarkers as these are expressed differentially in neurodegenerative conditions and might, intuitively, provide a more holistic insight into heterogenous pathologies represented in real-world, memory clinic settings (38). For instance, NFL has been shown to identify dementia in Down syndrome and amongst psychiatric conditions, and even between demented and non-demented Parkinson’s disease patients (39, 40). This gap might, in part, be due to the exclusion of comparisons of a selected biomarker with another biomarker like p-tau181 or plasma Aβ42/40. This biomarker selection trade-off was considered during the conceptualisation phase of the review, and was justified since, (a) p-tau217 has shown superior performance to other p-tau isoforms (8, 41–44), (b) plasma Aβ42/40 assays have shown limited robustness and diagnostic range (45, 46), and (c) an intention to keep biomarker selection consistent with the *Revised Criteria* was a major driving force for the current review.

A similar trade-off was also considered while excluding data from exclusively cognitively healthy, pre-clinical populations, or those from population-based- or community-cohorts. Importantly, as community- or population-cohort enrolment hinges on voluntary participation, often, at-risk groups are inherently excluded, and this limits generalisability in clinical populations. Yet, as much of clinical research has utilized data from cohorts and health registries, articles were included if the cohort mentioned at least partial recruitment from clinical services. Notably, through the screening process, it was observed that cohorts including minority- or non-White ethnicities recruited exclusively through community outreach initiatives and not through clinical services, suggesting a lack of healthcare utilization and access for ethnic minority groups.

Eleven studies represented patient populations from World-Bank designated high income countries (HICs): France, Germany, Italy, Netherlands, Portugal, Sweden, Spain, and USA, and two studies presented data from Chinese populations. Adjacently, only three included articles reported ethnicity, of which two reported >85% White participants. Geographical representation was also limited, with only one study being conducted in a non-High-income country. This limitation reflects disparities in resources and infrastructure, which perpetuate the use of clinical judgement alone, without biomarker-backed diagnoses, in resource-deficient settings (47, 48). This creates a paradox: while the use of blood biomarker tests might bridge this disparity, their validation requires data from the very populations that currently lack access to currently-approved, gold standard methods. Efforts to improve inclusion and ethnicity reporting in clinical cohorts must be emphasised in future research. In addition to ethnicity, many analyses in the included article failed to consider social determinants of health (SDoH), such as social contact, socioeconomic status, and discrimination, that have increasingly shown an association with dementia outcomes (49, 50). This omission also reduces the applicability and relevance of the above findings to historically marginalized groups. Additionally, it is important to note that while the review’s exclusive focus on cross-sectional studies in memory clinic settings provides increased relevance in routine clinical practice, it restricts the scope of BBM utility in earlier phases of the AD continuum, which has been widely demonstrated (43, 51).

Despite the growing body of evidence showcasing the effectiveness and advantages of plasma biomarkers, significant gaps in the literature persist, especially in terms of representation of real-world, diverse, clinical settings, and the absence of contextual variables. These issues need to especially considered in the context of blood biomarkers, given that they provide a highly accessible, globally scalable, and resource-effective alternative for current methods.

## Data Availability

All data produced in the present work are contained in the manuscript and supplementary materials.

---

**Supplementary Figure 1.**
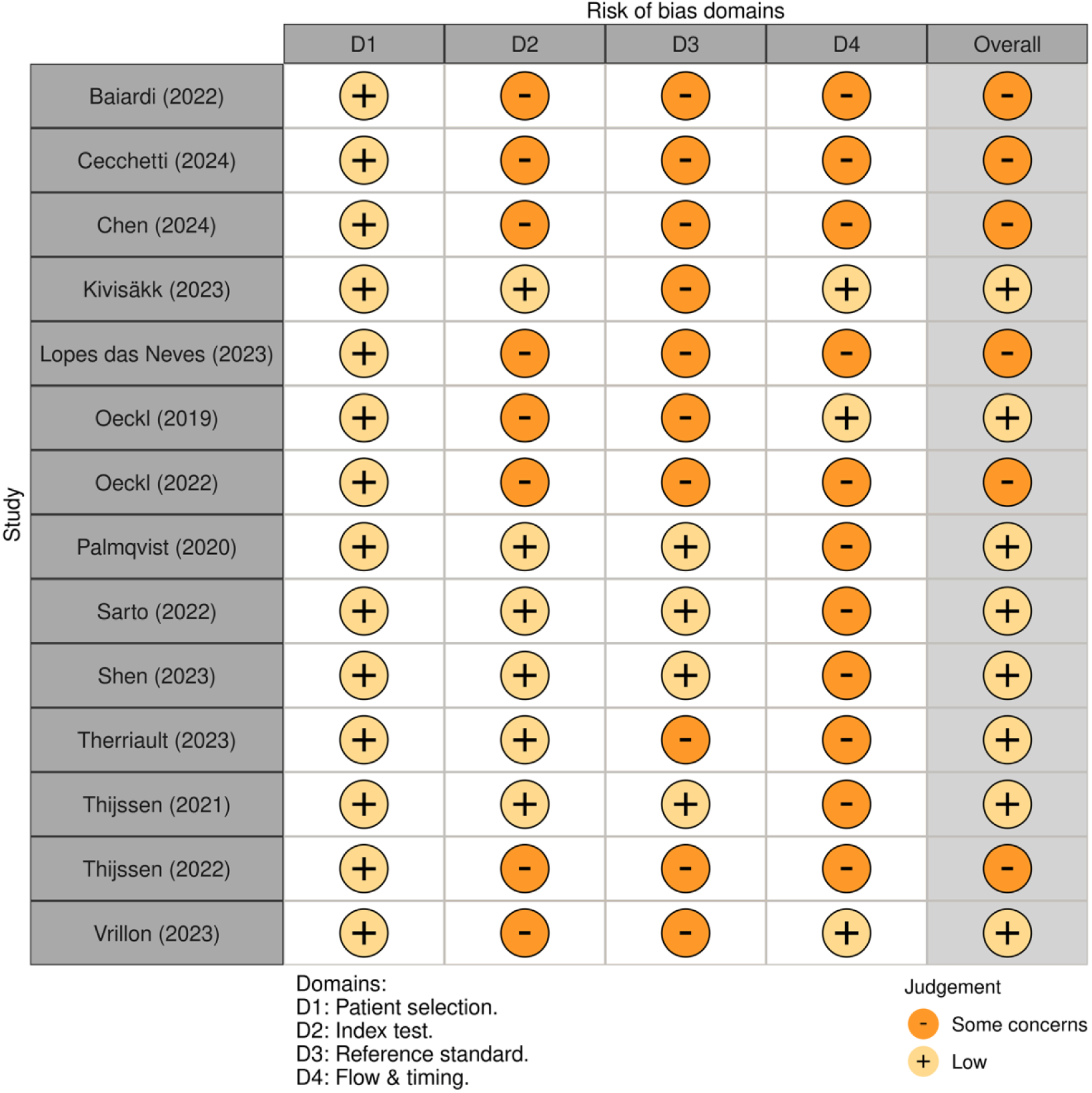
Risk of Bias assessment for selected studies using the QADAS-2 tool.

